# Mapping the Cerebral Burden of Status Epilepticus – Results from a Longitudinal MRI Study

**DOI:** 10.64898/2026.05.15.26353238

**Authors:** Bernardo Crespo Pimentel, Pilar Bosque-Varela, Lukas Machegger, Luca Panebianco, Jürgen Steinbacher, Johannes Pfaff, Fenglai Xiao, Markus Leitinger, Eugen Trinka, Giorgi Kuchukhidze

## Abstract

Status epilepticus (SE) is a neurological emergency with a mortality of up to 39% in population-based studies. Animal studies suggest that, beyond a critical timepoint (t2), SE induces neuronal injury exceeding what can be expected from the underlying aetiology, but in vivo evidence in humans is scarce. Thirty-six prospectively recruited individuals with SE with a mean age of 59 years underwent serial high-resolution T1-weighted brain MRI over a mean follow-up period of 5 months and were compared with 34 individuals with drug-resistant focal epilepsy and 36 propensity-score matched healthy controls. Cortical thickness and deep grey matter volumes were estimated and harmonised to account for scanner-related effects.

Longitudinal change was assessed using linear mixed-effects models adjusting for age at baseline, interscan interval and sex between groups. We further investigated the independent effects of SE duration, semiology and level of consciousness after mutual adjustment for each variable as well as aetiology, as well as longitudinal structural signatures associated with key locations of peri-ictal MRI abnormalities (PMA). SE was associated with pronounced bilateral hippocampal atrophy in comparison to normal aging and drug-resistant epilepsy, alongside volume increase in several deep grey matter nuclei as well as a trend for cortical thinning of medial brain structures. SE duration was the strongest independent driver of brain change, producing widespread cortical thinning and bilateral hippocampal atrophy. A convulsive semiology was independently associated with accelerated medial temporal cortical thinning and bilateral hippocampal volume loss when compared with non-convulsive (NCSE) and other prominent-motor SE, while reduced consciousness predicted faster thinning of medial frontoparietal cortex. PMA was associated with distinct longitudinal trajectories of subcortical volume, with pulvinar and hippocampus involvement predicting thalamic and hippocampal atrophy patterns. According to our findings, a single episode of SE was therefore associated with a measurable structural imprint in the brain that evolved over months beyond what would be expected for aetiology alone. Atrophy trajectories highlighted the vulnerability of the hippocampus and other limbic structures during the peri-ictal state. A long SE duration, together with convulsive semiology and impaired consciousness, independently amplified this damage. These findings corroborate the timepoint-based (t2) concept of SE and reinforce the clinical imperative of rapid seizure termination to contain long-term structural brain injury.

## Introduction

More than fifty years ago, it was proposed that seizures occurring in early life may lead to mesial temporal sclerosis.^1^ However, this causal relationship still remains poorly understood. Despite decades of progress in epilepsy research, an important gap remains in our understanding of the long-term consequences of prolonged seizures on the human brain.

Status epilepticus (SE) is a neurological emergency associated with a mortality rate of up to 39%.^2,3^ It results from failure of mechanisms responsible for seizure termination or from the initiation of mechanisms which lead to abnormally prolonged seizures (after timepoint t1).^4^ Evidence arising from animal models supports the concept that SE can induce irreversible neuronal injury (after timepoint t2).^4–7^ Based on this, a timepoint of t2 at 30 min was proposed in human convulsive SE (CSE) and 60 min in non-convulsive SE (NCSE), whereas evidence regarding SE with prominent motor symptoms (PM-SE), other than CSE, is limited. In humans, in vivo neuroimaging evidence of SE-induced brain damage as surrogate marker for timepoint t2 has accumulated over recent years, although existing studies have often been limited by small sample sizes, lack of quantitative methodologies and longitudinal designs.^8–13^

According to the “impact of burden model” of SE, ictal-related brain injury adds to the tissue damage caused by the underlying aetiology and comprises all metabolic, structural, genetic and network-related effects of SE on the brain^14,15^. This added burden may be mediated by seizure-induced disruption of the ionic milieu, which promotes glutamate release, calcium influx into the cells, cytotoxic oedema and ultimately neuronal death.^16,17^ Peri-ictal MRI abnormalities (PMA) such as compensatory hyperperfusion, vasogenic and cytotoxic oedema have been reported in about 46% of individuals with SE.^12^ PMA in DWI/FLAIR imaging are considered to represent a neuroimaging correlate of possible neuronal injury after timepoint t2, and reflect the toll of prolonged seizure activity on neuronal homeostasis, potentially leading to permanent structural and functional damage in brain networks.^12,18^ Gene expression changes subsequently occur in seizure-damaged brain tissue, driving long-term network remodelling.^19,20^ The occurrence of long-term cognitive impairment following SE further supports an underlying convergent neurodegenerative process.^21,22^

In this retrospective analysis of a prospectively recruited SE cohort, we used serial MRI to track structural brain changes over time, following SE. We aimed to map brain changes in SE via established MRI-markers of neurodegeneration and investigate whether PMA location is associated with divergent atrophy trajectories. Building on previous reports on the same data^11^, we hypothesized that SE is associated with accelerated brain atrophy as compared to normal aging and drug-resistant focal epilepsy.^23,24^ Furthermore, we propose that the cerebral burden of SE is independently modulated by clinical features such as level of consciousness, semiology and duration of SE, beyond the effect of the underlying aetiology.

## Materials and methods

### Participants

#### Status epilepticus

Five hundred and ninety adult patients with SE were prospectively recruited between February 2019 and April 2024 at the Christian-Doppler University Hospital, Paracelsus Medical University, in Salzburg, Austria. The diagnosis of SE was based on the International League Against Epilepsy (ILAE) criteria; the Salzburg Consensus EEG Criteria and the definitions proposed in the American Clinical Neurophysiological Society Terminology 2021 were employed for diagnosing NCSE.^4,25–28^ In 375 (64%) of participants with SE, MRI was performed within 48 hours of onset of SE. Fifty-seven (15%) participants aged 21 to 88 years exhibited PMA and underwent follow-up imaging at least four weeks after the baseline imaging. We excluded individuals with underlying conditions potentially associated with neurodegeneration, including dementia (N=3) and hypoxic brain injury (N=1) due to cardiac or respiratory arrest and mitochondrial diseases (N=2). We further excluded individuals with brain surgery or another SE between MRI scans, individuals with at least one MRI scan with insufficient quality or lack of tridimensional T1-weighted sequences (N=11) and individuals with extensive brain lesions (N=4). Our complete SE MRI protocol is described in the Supplementary Information 1. MRI scans were assessed for the occurrence and location of PMA on diffusion-weighted imaging (DWI), fluid-attenuated inversion recovery (FLAIR) and arterial spin labeling (ASL) by two experts with extensive experience in neuroimaging (Lu.M. and G.K.). The criteria to classify an MRI lesion as PMA are described in Supplementary Information 2. In analogy to a previously published report on the same cohort, duration of SE was defined as the known or presumed time from symptom onset to clinical or EEG cessation of SE. Aetiology was classified as (1) acute symptomatic, (2) remote symptomatic, (3), progressive symptomatic and (4) unknown.^4^ No patients had defined electroclinical syndromes, such as idiopathic generalized epilepsy. We divided patients based on the level of consciousness into two groups: (1) those who were alert or somnolent, and (2) those in a stupor or coma. Consciousness was assessed clinically before treatment with benzodiazepines and first-line ASMs.

#### Drug-resistant focal epilepsy

To control for longitudinal brain structure change associated with chronic difficult-to-treat epilepsy, we included a cohort of individuals with drug-resistant focal epilepsy according to the ILAE definition^29^. We identified 55 individuals with focal epilepsy who underwent a cerebral MRI between 2010 and 2013 from our video-EEG monitoring epilepsy database and had serial high-resolution T1-weighted scans at least two months apart (see Supplementary Information 1 for detailed MRI protocol). We excluded drug-responsive individuals (N=7), individuals who underwent surgery or had SE up until the timepoint of the last MRI scan (N=3), as well as datasets with insufficient image quality in any of the scans (N=8) or with conspicuous brain lesions (N=3). Selected individuals were aged 18-77 years. Presumed epilepsy location and lateralisation was based on clinical, electrophysiological and neuroimaging data. Unclear lateralisation or localisation was classified as undetermined.

#### Healthy controls

We included a cohort of healthy controls aged 31-80 years old with at least two high-resolution T1-weighted brain MRI scans more than two months apart. Data were collected from the Parkinson’s Progression Markers Initiative (PPMI) database (www.ppmi-info.org/access-data-specimens/download-data), a publicly available anonymized MRI database. The healthy control cohort is described in Supplementary Information 3. One individual was excluded due to preprocessing errors. Propensity score matching was performed in healthy controls to participants with SE 1:1 using a nearest-neighbour algorithm with replacement based on age and sex.

All study procedures were performed in accordance with the Declaration of Helsinki. The study was approved by the ethics committee of the state of Salzburg (Ethikkommission für das Bundesland Salzburg).

### MRI Preprocessing

We estimated vertex-wise and general cortical thickness through the validated Computation Anatomy Tool (CAT12) using an inverse-consistent longitudinal surface registration approach.^30^ We used the Hipposeg algorithm for accurate measurement of hippocampal volume.^31^ Hipposeg is an open-source, multiatlas-based, previously validated hippocampal segmentation algorithm that is robust to hippocampal morphologic alterations, including atrophy.^32^ It has demonstrated superior delineation of diseased hippocampi as compared to other automated segmentation methods.^32^ Volumes of other subcortical structures and total intracranial volume were measured using the Geodesic Information Flow algorithm.^33^ Both algorithms are accessible on NiftyWeb (http://cmictig.cs.ucl.ac.uk/niftyweb).

### Statistical Analysis

Demographical and clinical data were analysed with R studio (version 2023.03.0+386). Categorical variables are displayed as numbers and percentage and were analysed with Fisher’s exact test. Continuous variables are presented as mean ± SD (standard deviation) and were analysed with two-sample t tests for two-group comparisons and one-way analysis of variance (ANOVA) for three-group comparisons. Post-hoc tests were performed using Bonferroni-corrected pairwise comparisons. Missing data was handled by pairwise deletions.

#### Harmonisation of scanner-related effects

MRI-derived cortical thickness and subcortical volume measures were harmonised using the longitudinal ComBat Harmonisation Tool.^34^ For group comparisons (i.e., SE vs. healthy controls or focal epilepsy), we modelled age, group, time, and the group-by-time interaction as biological covariates to be preserved, with a subject-specific random intercept to account for repeated measurements within individuals. For the individual effects of clinical variables (e.g., SE duration, semiology, aetiology, etc.), we subset the data to the SE cohort and harmonised with the same mutually adjusted longitudinal model employed downstream, including a subject-specific random intercept.

#### Modelling of cortical thickness and subcortical volumes

Harmonised cortical thickness measurements were analysed with BrainStat within MATLAB (https://brainstat.readthedocs.io/en/master/) using a vertex-wise approach, whereas harmonised subcortical volumes were analysed with R studio (version 2023.03.0+386), using a region of interest (ROI)-wise approach.^35^ Global measures of brain structure including white matter volume (WMV) and global grey matter volume (GMV) were derived from the tissue segmentation results of CAT12. Cortical thickness and volume estimates were fitted into linear mixed-effect models to account for repeated measurements per subject with irregular measurement intervals.

#### Longitudinal brain structural change between groups

To investigate group-differences on the rate of cortical thickness (vertex-wise) or rate of subcortical volumes change over time, we tested for a main effect of the interaction between the grouping variable (e.g., “SE” and “focal epilepsy”) and interval between baseline and index MRI scan in months (with baseline = 0 months). We corrected for a random effect of subject and fixed effect of group allocation, time interval in months and sex. We report cortical thickness findings at *P*<0.05 both uncorrected and corrected for multiple comparisons via random field theory (RFT) at family-wise error (FWE) <0.05. Findings on volumetry of subcortical structures are reported after multiple comparisons through false-discovery rate (FDR) at *P*<0.05. To increase sensitivity to seizure-related structural alterations and assess effects relative to the hemisphere of ictal onset, all datasets from participants with right-sided SE onset or focal epilepsy location were mirrored across the midline. This ensured that the hemisphere of ictal onset was consistently represented as the left (ipsilateral) hemisphere in all analyses, with the right hemisphere representing the contralateral side. Datasets from participants with unknown side of SE or focal epilepsy were left unmirrored.

#### Modulators of longitudinal structural change following SE

We tested vertex- and ROI-wise whether each predictor variable (semiology, consciousness and duration of SE) independently explained differences in the rate of longitudinal cortical thickness or subcortical volume change, beyond the corresponding time-dependent effects of aetiology (as categorical variable with 4 levels: acute, remote, progressive and unknown)^4^, the other variables, age at baseline and sex. Omnibus effects on longitudinal change were assessed with nested-model F tests comparing a full model including the corresponding timepoint by predictor interaction against an otherwise identical reduced model omitting that interaction. To enable valid omnibus inference, models were fitted as fixed-effects with subject-specific intercepts. Multiple-comparison control was performed using RFT (FWE *P*<0.05) for cortical thickness and via FDR for subcortical volume estimates. To investigate directionality of changes (i.e., more or less atrophy over time), we performed follow-up analyses for pairwise comparisons between different levels of the study variables (e.g. for semiology, CSE vs. NCSE and CSE vs. other PM-SE) using mutually adjusted pairwise slope contrasts while additionally correcting for aetiology and age at baseline.

### Location of DWI/FLAIR PMA and longitudinal structural brain change

We examined the atrophy trajectories according to DWI/FLAIR PMA status in three key regions: the cortex, pulvinar nucleus of the thalamus and the hippocampus. These regions were selected based on prior work in this cohort demonstrating their specific involvement in SE.^36^ Analogous to the group comparisons described above, vertex- and ROI-wise linear mixed models were performed with PMA status at the index location as the grouping variable. Models included fixed effects the interaction of timepoint and both SE aetiology and SE duration as well as age at baseline and a subject-level random effect. Multiple-comparison control was performed using RFT (FWE *P*<0.05) for cortical thickness and via FDR for subcortical volume estimates. Individuals with PMA involving multiple regions were included in each relevant subgroup, such that participants could contribute to more than one location-specific analysis.

## Results

### Demographic and clinical characteristics

We assessed 251 longitudinal high-resolution T1-weighted MRI datasets from a total of 106 individuals. After exclusion of study participants (N=43) and propensity-score matching, our analysis included 36 individuals with SE, 34 individuals with drug-resistant focal epilepsy and 36 healthy controls. Fourteen (38%) individuals with SE had 4 longitudinal MRI measurements, 11 (31%) obtained 3 MRIs and 11 (31%) obtained 2 MRIs, at different timepoints after SE. The group of healthy controls was comparable to the SE group concerning age at baseline imaging and sex. However, the SE cohort was older at baseline imaging than the drug-resistant focal epilepsy group [59 (17) vs. 31 (11) years, *P*<0.001], and time between baseline and last scan differed significantly between the SE group and both the focal epilepsy [150 (218) vs. 391 (412) days, *P*<0.001] and healthy cohorts [150 (218) vs. 384 (99) days, *P*<0.001]. Among the focal epilepsy patients, 16 (47%) were classified as temporal lobe epilepsy, 4 (12%) as frontal lobe epilepsy, 4 (12%) as parietal lobe epilepsy, 4 (12%) as unknown localisation, 3 (8.8%) as occipital lobe epilepsy, and 1 (2.9%) as multifocal epilepsy. Demographic characteristics of our research groups are displayed in Table 1.

**Table 1.**
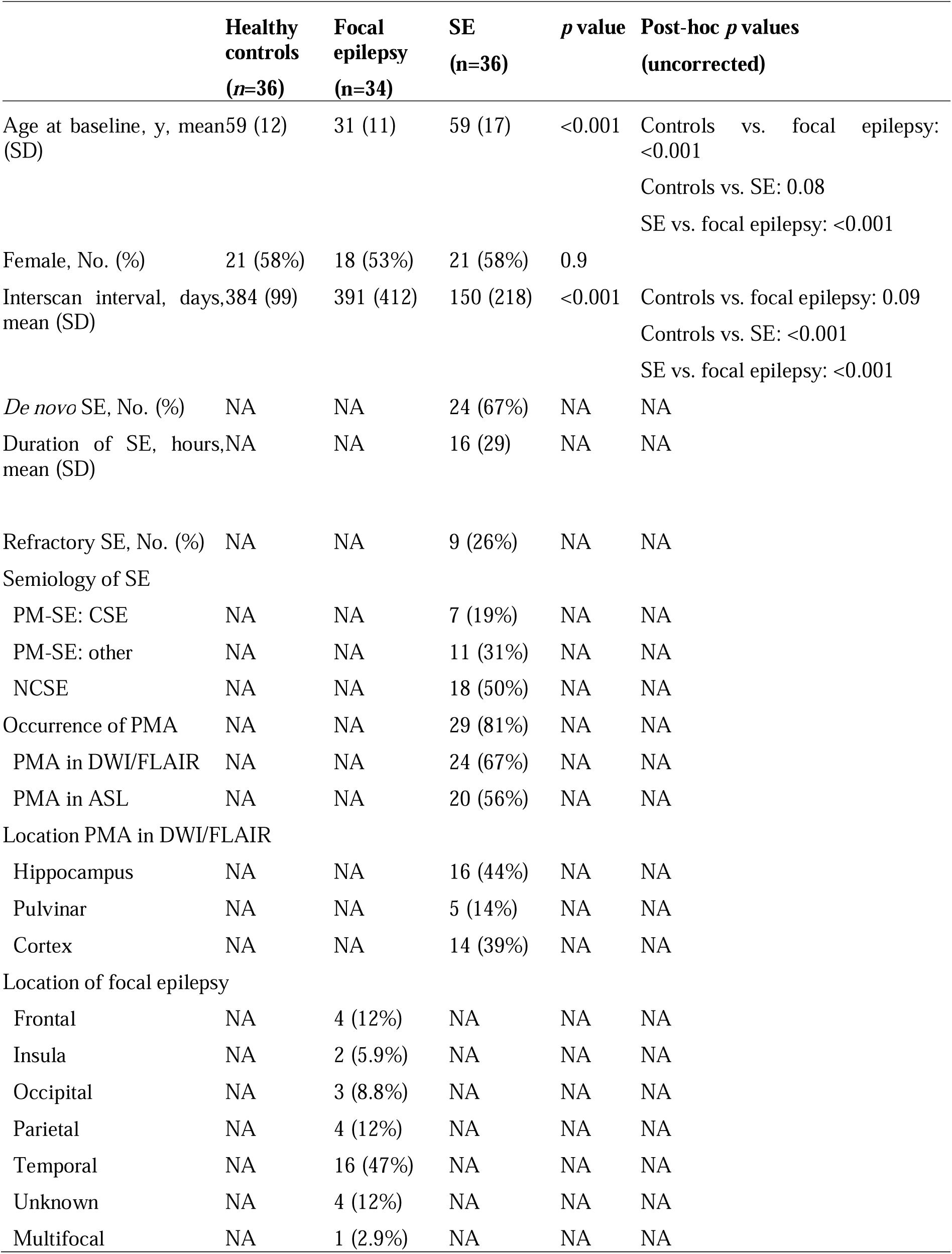
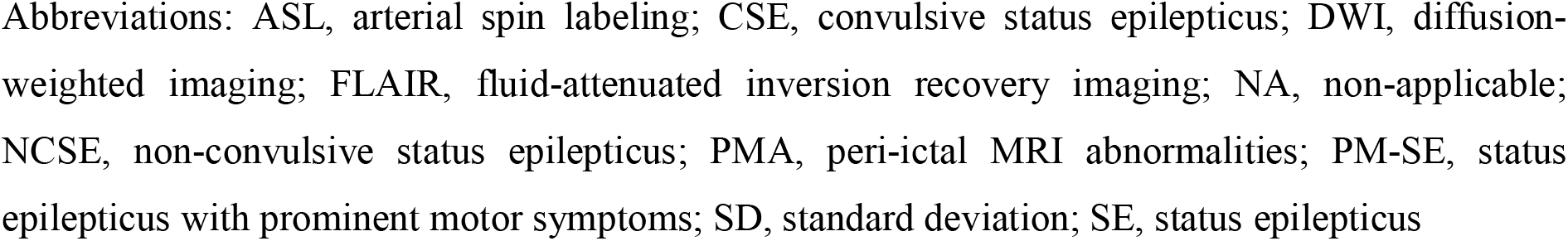
Baseline demographic and clinical characteristics.

Eighteen (49%) of participants with SE exhibited prominent motor features (PM-SE), of whom 7 (19%) individuals presented bilateral tonic-clonic activity and were therefore classified as CSE. Eighteen (50%) of SE participants were classified as NCSE. An acute symptomatic aetiology was accounted for in 16 (44%) of SE participants, whilst 5 (14%) had a remote symptomatic and 7 (19%) a progressive symptomatic cause. Aetiology remained unknown in 8 (22%) individuals with SE. Nine participants (26%) were resistant to benzodiazepine and first-line anti-seizure medication (ASM) and were as such categorised as refractory SE (RSE). Twenty-five individuals (69%) with SE presented clinically in an alert or somnolent state, whereas 11 participants (31%) were in stupor or coma. PMA was observed on baseline imaging in 29 (81%) individuals with SE. From these, 24 individuals (67%) exhibited PMA in the DWI and/or FLAIR sequences. Details regarding PMA location are found in Table 1, whereas other clinical characteristics of our SE group are listed in Table 2.

**Table 2.** Baseline characteristics of the SE group.

|  | All SE<br>(n=36) | CSE<br>(n=7) | Other PM-SE<br>(n=11) | NCSE<br>(n=18) | p value |
| --- | --- | --- | --- | --- | --- |
| Age at baseline, years, mean (SD) | 59 (17) | 57 (22) | 60 (15) | 58 (17) | .89 |
| Female, No. (%) | 21 (58%) | 3 (43%) | 8 (73%) | 10 (56%) | >0.99 |
| De novo SE, No. (%) | 24 (67%) | 5 (71%) | 7 (64%) | 12 (67%) | >0.99 |
| Duration of SE, hours, mean (SD) | 16 (29) | 2 (2) | 10 (22) | 25 (34) | .06 |
| Refractory SE, No. (%) | 9 (26%) | 0 (0%) | 6 (35%) | 3 (27%) | .20 |
| Aetiology, No. (%) |  |  |  |  |  |
| Acute symptomatic | 16 (44%) | 2 (29%) | 5 (45%) | 8 (44%) | NA |
| Remote symptomatic | 5 (14%) | 2 (29%) | 1 (9.1%) | 2 (11%) | NA |
| Progressive symptomatic | 7 (19%) | 1 (14%) | 4 (36%) | 3 (17%) | NA |
| Unknown | 8 (22%) | 2 (29%) | 1 (9.1%) | 5 (28%) | NA |
| Outcome |  |  |  |  |  |
| Days spent in ICU, mean (SD) | 6 (9) | 10 (16) | 5 (6) | 5 (7) | .54 |
| Admission to ICU, No. (%) | 21 (60%) | 5 (71%) | 6 (55%) | 10 (59%) | >0.99 |
| EMSE, mean (SD) | 51 (30) | 31 (17) | 59 (34) | 61 (28) | .08 |
| STESS, mean (SD) | 2.66 (1.43) | 3.43 (1.51) | 2.18 (1.25) | 2.65 (1.46) | .97 |

Abbreviations: CSE, convulsive status epilepticus; EMSE, epidemiology-based mortality score in Status epilepticus; NA, non-applicable; NCSE, non-convulsive status epilepticus; ICU, intensive care unit; PMA, peri-ictal MRI abnormalities; PM-SE, status epilepticus with prominent motor symptoms; SD, standard deviation; SE, status epilepticus; STESS, Status epilepticus severity score

#### Longitudinal changes of brain structure following SE

WMV in SE declined at a rate of 0.81 ml/month (*P*_FDR_=0.04), faster than in focal epilepsy (+0.08 ml/month; *P*_FDR_=0.05) and healthy controls (+0.121 ml/month; *P*_FDR_=0.05), whereas GMV alterations were not significant, as depicted in Supplementary Figure 1A. Whole-brain plots in Supplementary Figure 1B show a tendency for bilateral thinning in the parahippocampal, bilateral posterior cingulate gyrus, right anterior cingulate gyrus and left superior temporal gyrus in the SE group, although not reaching statistical significance after RFT correction. In the hippocampus we observed a significant bilateral volume decline reaching -120.25 mm^3^/month on the left side (*P*_FDR_<0.05), whereas other subcortical grey structures gained significant volume, including bilateral thalamus (left: 100 mm^3^/month, *P*_FDR_<0.05; right: 109.26 mm^3^/month, *P*_FDR_<0.05), bilateral putamen (left: 95.83 mm^3^/month, *P*_FDR_<0.05; right: 83.17 mm^3^/month, *P*_FDR_<0.05), bilateral caudate nuclei (left: 58.04 mm^3^/month, *P*_FDR_<0.05; right: 72.81 mm^3^/month, *P*_FDR_<0.05), bilateral accumbens nuclei (left: 12.14 mm^3^/month, *P*_FDR_<0.05; right: 12.14 mm^3^/month, *P*_FDR_<0.05), among others. Please refer to Supplementary Tables 1 and 2 for detailed results.

#### SE is associated with mild cortical thinning and divergent subcortical volume change

Major group comparisons yielded overall a tendency for cortical thinning in SE in medial brain regions. Against healthy controls, Figure 2A shows clusters in the cingulate gyrus and temporal lobes (e.g., right cingulate gyrus T=-2.03, *P_uncorrected_* <0.05, right superior temporal lobe T=-2.59, *P_uncorrected_* <0.01). Similar areas also presented significant thinning as compared with focal epilepsy (e.g., right posterior cingulate T=-3.7, *P_uncorrected_*<0.001 and left superior temporal gyrus T=-3.15, *P_uncorrected_*<0.001). These significant findings lateralised to the ipsilateral side of SE onset and seizure activity in focal epilepsy (e.g., ipsilateral posterior cingulate cortex T=-3.7, *P_uncorrected_*<0.001, ipsilateral superior temporal gyrus T=-2.76, *P_uncorrected_*<0.01), as displayed in Figure 2B. None of these clusters survived multiple comparison correction via RFT. Detailed results are listed in Supplementary Table 3.

**Figure 1.**
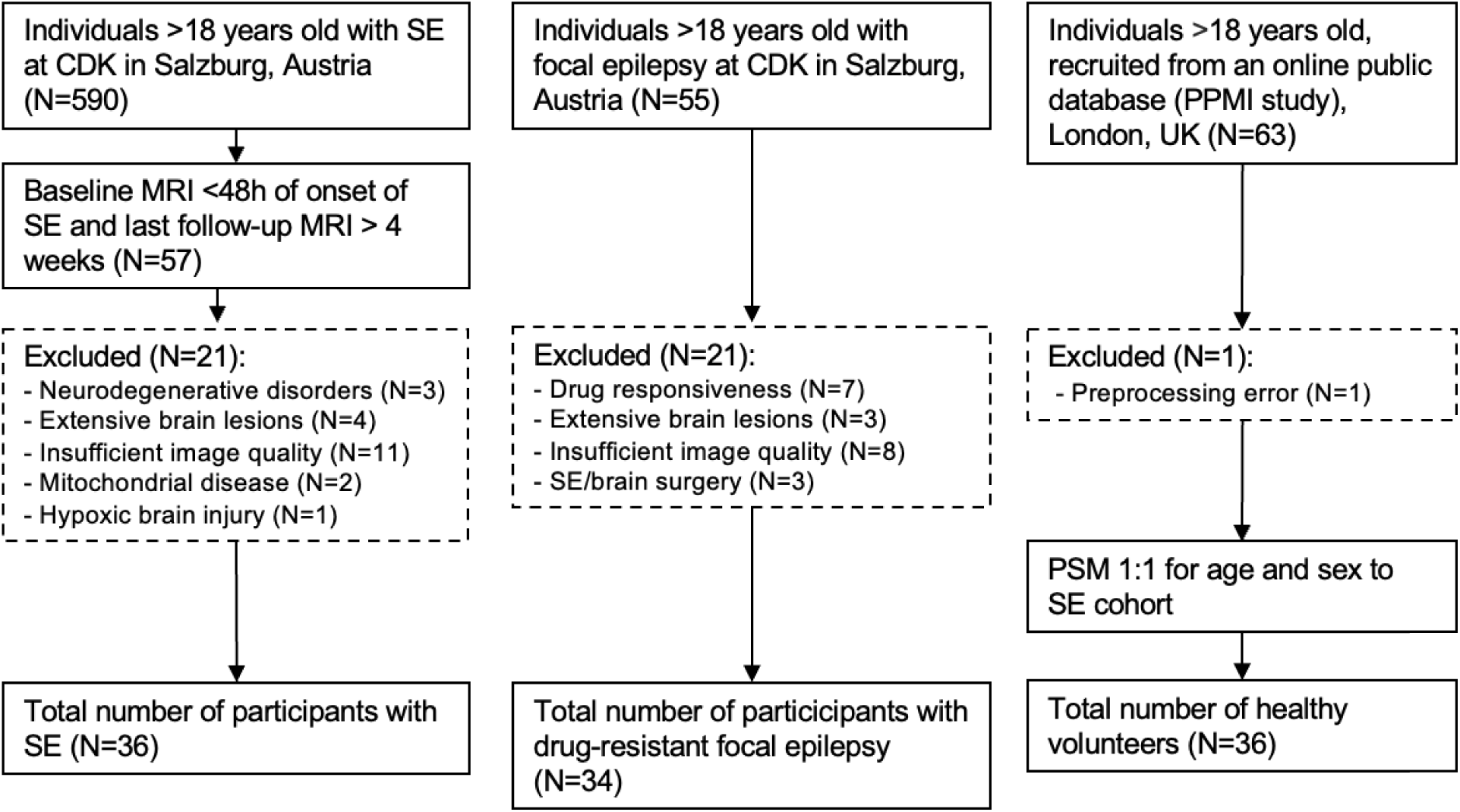
Study flowchart. Abbreviations: CDK, Christian-Doppler University Hospital; PSM, propensity-score matching; SE, status epilepticus

**Figure 2.**
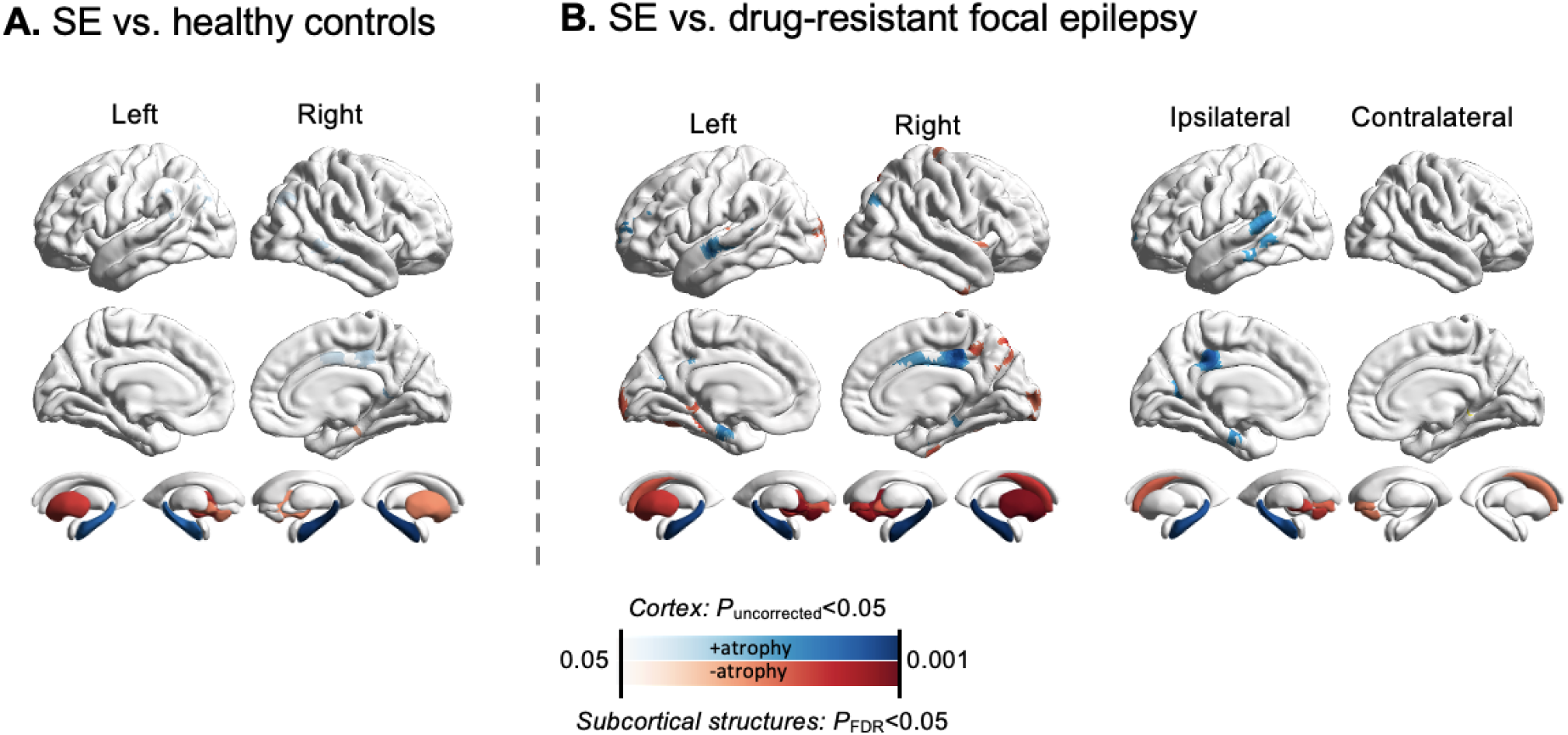
Cortical thinning and distinct subcortical volume trajectories in SE compared to healthy controls and focal epilepsy. Brain-wide plots depict SE-associated accelerated atrophy or conversely less atrophy, compared with (A) normal aging and (B) drug-resistant focal epilepsy. For all group comparisons, no cortical clusters survived RFT correction at FWE <0.05, therefore clusters with uncorrected *P* values <0.05 are shown. For subcortical volume estimates, results are restricted to structures with FDR correction at *P*<0.05. The maps suggest trends towards thinning of the cingulate and lateral temporal cortex in SE and show significant divergent volume longitudinal trajectories in deep grey matter structures, including consistent hippocampal atrophy in comparison to both healthy controls and drug-resistant focal epilepsy. In (B), we provide a comparison between SE and drug-resistant focal epilepsy with respect to the side of SE onset or, in focal epilepsy, seizure focus. This suggests lateralised accelerated atrophy and subcortical volume change on the side of SE onset. Abbreviations: FDR, false-discovery rate; FWE, family-wise error; RFT, random field theory; SE, status epilepticus

Accelerated hippocampal volume decline in SE was a transversal finding in all group comparisons, against healthy controls (left -73.69 mm^3^/month faster, *P*_FDR_ <0.001, right - 76.10 mm^3^/month faster, *P*_FDR_ <0.001) and drug-resistant focal epilepsy (left -83.17 mm^3^/month faster, *P*_FDR_ <0.001; right -87.40 mm^3^/month faster, *P*_FDR_ <0.001). The basal ganglia showed mild volume gains after SE when compared to age-dependent changes (e.g., left pallidum: 36.14 mm^3^/month faster, *P*_FDR_ <0.05, right putamen 41.60 mm^3^/month faster, *P*_FDR_ <0.05), and focal epilepsy (e.g. left pallidum: 34.11 mm^3^/month faster, *P*_FDR_ <0.01 and left putamen: 46.43 mm^3^/month faster, *P*_FDR_<0.001). When analysing with respect to side of SE onset (Figure 2B), the ipsilateral hippocampus relative to the side of SE showed the fastest volume decline in all group comparisons (−126.07 mm^3^/month faster, *P*_FDR_<0.001), whereas the contralateral hippocampus did not show any significant volume dynamic (−34.66 mm^3^/month faster, *P*_FDR_ =0.52). Furthermore, as significant volume gains were observed in the putamen bilaterally (ipsilateral: 48.51 mm^3^/month faster, *P*_FDR_ <0.01; contralateral: 127 mm^3^/month faster, *P*_FDR_<0.05), accumbens nucleus (ipsilateral: 7.91 mm^3^/month faster, *P*_FDR_ <0.01; contralateral: 18.75 mm^3^/month faster, *P*_FDR_ <0.05) and caudate nucleus (ipsilateral 71.94 mm^3^/month faster, *P*_FDR_ <0.05; contralateral 94.94 mm^3^/month faster, *P*_FDR_ <0.05). Results are displayed in Figure 2A and 2B and listed in detail in Supplementary Figure 3. Results regarding longitudinal brain structure changes in refractory and non-refractory SE are presented in Supplementary Figure 1.

#### Independent contributions of SE semiology, duration and consciousness level

We employed omnibus nested F-tests to show the independent effects of aetiology on the longitudinal variation of cortical thickness and subcortical volumetry. Aetiology showed widespread effects on cortical morphology variation but none over subcortical structures, including hippocampus. These results are displayed in Supplementary Figure 2.

Concerning level of consciousness on admission, omnibus nested F-tests showed effects on longitudinal variation of cortical thickness restricted to the ipsilateral medial precentral, posterior cingulate and superior frontal gyrus (F=21,81, *P*_FWE_<0.05) as well as contralaterally to a cluster involving the precuneus, paracentral and posterior cingulate gyrus, among others (F=23,47, *P*_FWE_<0.01). Across these significant clusters, a stuporous or comatose state was associated with accelerated cortical thinning of 0.20 mm/month (*P*=0.01) in comparison to an alert/somnolent state (Figure 3A).

**Figure 3.**
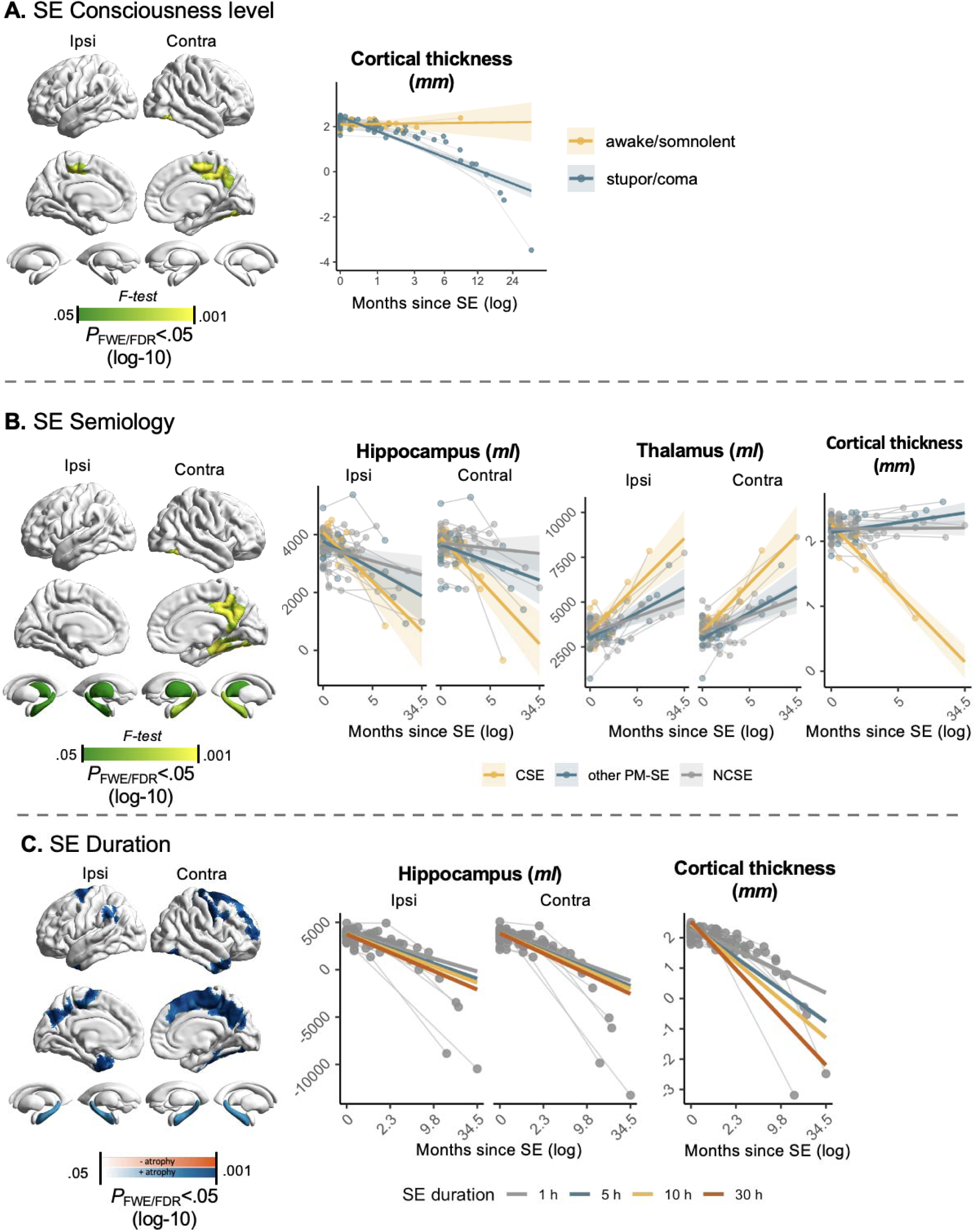
Independent contribution of clinical variables to progressive structural change after SE. Whole-brain maps depict regions in which (A) level of consciousness, (B) semiology, and (C) SE duration were independently associated with altered rates of longitudinal cortical thickness or subcortical volume change after mutual adjustment for one another, as well as for aetiology, age at baseline and sex. Cortical effects were identified by using omnibus tests of the relevant moderator by time variable term in models and are shown at RFT-corrected FWE *P*<0.05. Subcortical effects were tested in analogous mutually adjusted mixed-effects models and are shown at FDR-corrected *P*<0.05. For SE duration, effect maps are signed to indicate the direction of the association with longitudinal change. The accompanying scatter plots show nuisance-residualised longitudinal values from the full mutually adjusted model, preserving time and the moderator of interest and therefore reflecting associations adjusted for the remaining covariates. Abbreviations: FDR, false-discovery rate; FWE, family-wise error; RFT, random field theory; SE, status epilepticus

Comparable semiology-related effects were observed. Figure 3B shows significant independent effects of SE semiology in the contralateral fusiform, parahippocampal, lingual, entorhinal and inferior temporal gyrus (F=20.5 *P*_FWE_<0.0001), as well as in the precuneus and posterior cingulate gyrus (F=9.86; *P*_FWE_<0.01). Here, CSE was associated with a faster cortical thinning compared to both NCSE and other PM-SE (difference 0.19 and 0.20 mm/month, respectively; *P*<0.01). Both hippocampi also showed a significant accelerated volume decline associated with CSE as compared to NCSE (ipsilateral: -315.25 mm^3^/month faster, *P*<0.01; contralateral: -434 mm^3^/month faster, *P*<0.01). CSE was associated with a significant volume decline as compared to other PM-SE (ipsilateral: -234.18 mm^3^/month faster, *P*<0.01; contralateral: -345.30 mm^3^/month faster, *P*<0.01).

Lastly, SE longer duration was independently associated with widespread cortical thinning involving ipsilateral areas of the frontal, parietal and temporal lobes as well as contralateral medial frontal lobe, precuneus and temporal pole, all surviving RFT correction at FWE<0.001 (Figure 3C). The biggest cluster comprised the contralateral superior frontal, precentral, paracentral, middle frontal, posterior cingulate and superior parietal gyrus, among other regions (T=-21,75; *P*_FWE_<0.0001). The highest F-value was reported in a cluster comprising the ipsilateral precuneus, cuneus and superior temporal gyrus (T=-33,37; *P*_FWE_<0.0001). In these regions, a SE with a duration of 1 hour was associated with a mean cortical thinning of 0.12 mm/month (*P*=0.010), whereas 5 hours was associated with 0.19 mm/month (*P*<0.001). Regarding subcortical structures, duration of SE only showed an independent negative effect on the ipsi- and contralateral hippocampi (T=-9,10; *P*_FDR_<0.01 and T=-10,10; *P*_FDR_<0.05, respectively). Detailed results are listed in Supplementary Table 4. The mutually adjusted additional effect of aetiology on the longitudinal variation of cortical thickness and subcortical structures is displayed in Supplementary Figure 2.

#### PMA location is associated with divergent trajectories of structure change

Of all 36 individuals with SE, 16 (44%) exhibited DWI/FLAIR PMA involving the hippocampus, including one individual with bilateral involvement; 5 (14%) showed involvement of the pulvinar nucleus of the thalamus, all unilateral, from which 3 also exhibited hippocampus PMA; and 17 (47%) exhibited cortical involvement, including one with bilateral involvement. Among participants showing cortical PMA, 2 patients also had hippocampal PMA, and 2 had pulvinar PMA. Occurrence of PMA in the hippocampus in DWI/FLAIR imaging was independently associated with a significant volume decline of the ipsilateral hippocampus (-145.66 mm^3^/month, *P_FDR_*<0.01), ipsilateral thalamus (-160.36 mm^3^/month, *P_FDR_*<0.01) and ipsilateral amygdala (-107.83 mm^3^/month, *P_FDR_*<0.05). Pulvinar PMA was associated with volume decline of the ipsilateral accumbens nucleus (-14.21 mm^3^/month, *P_FDR_*<0.001), ipsilateral and contralateral amygdala (-50.10 mm^3^/month, *P_FDR_*<0.05 and -37.08 mm^3^/month, *P_FDR_*<0.01, respectively) and ipsilateral and contralateral thalamus (-82.65 mm^3^/month, *P_FDR_*<0.05 and -110.48 mm^3^/month, *P_FDR_*<0.01, respectively).

Cortical PMA was associated with accelerated volume decline in the ipsilateral hippocampus (-97.62 mm^3^/month, *P_FDR_*=0.01), as well as volume increases in the ipsilateral caudate nucleus (67.56 mm^3^/month, *P_FDR_*<0.05), contralateral accumbens nucleus (20.51 mm^3^/month, *P_FDR_*<0.05), contralateral amygdala (21.12 mm^3^/month, *P_FDR_*<0.05), contralateral caudate nucleus (109.19 mm^3^/month, *P_FDR_*<0.001), contralateral putamen (120.62 mm^3^/month, *P_FDR_*<0.05) and contralateral thalamus (86.77 mm^3^/month, *P_FDR_*<0.05). These results are displayed in Figure 4 and listed in detail in Supplementary Table 5.

**Figure 4.**
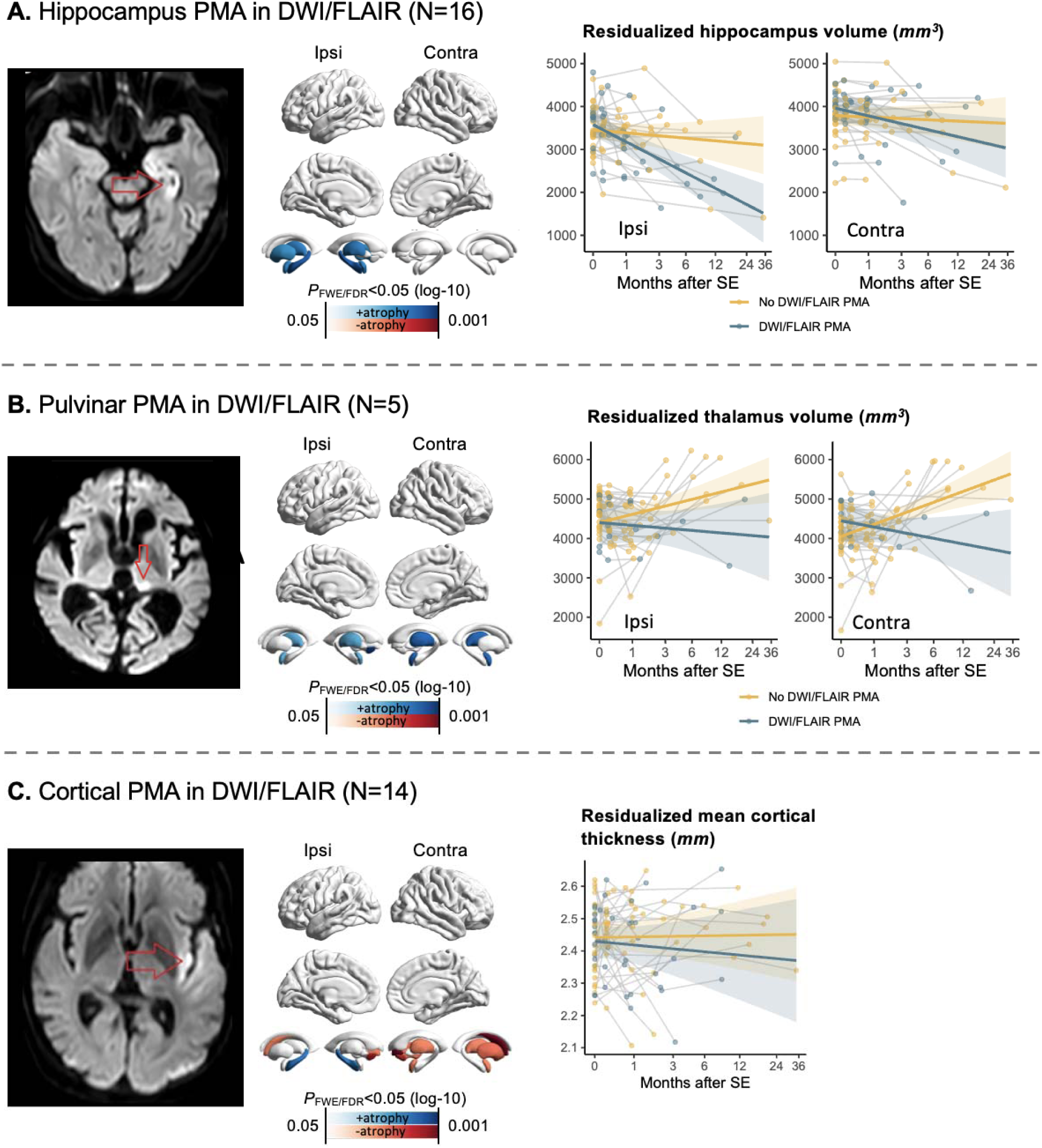
Location of PMA is associated with distinct subcortical volume trajectories. Whole-brain maps show regions in which the presence of PMA in DWI/FLAIR imaging in the (A) hippocampus, (B) pulvinar nucleus of the thalamus, and (C) cortex were independently associated with altered rates of longitudinal cortical thickness or subcortical volume change, after adjustment for the effects of aetiology, SE duration, as well as age at baseline and sex. No significant cortical clusters survived RFT correction at P_FWE_ <0.05, whereas PMA at different locations was associated with distinct subcortical volume trajectories. The accompanying scatter plots show nuisance-residualised longitudinal values from the model, preserving time and the moderator of interest. Because thalamic nuclei were not segmented separately, the whole thalamic volume is shown as a proxy of pulvinar volume.

## Discussion

In this longitudinal MRI cohort study, we estimated markers of neurodegeneration in a well-characterised cohort of 36 individuals with SE and tracked longitudinal trajectories of brain structure dynamics over a mean follow-up of 5 months. Brain changes after SE are complex, with trends to progressive cortical thinning as compared to normal aging and drug-resistant focal epilepsy, and more significant effects in the deep grey matter nuclei including hippocampi. We used statistical modelling to disentangle the contribution of SE aetiology on the rate of brain change over time from ictal-driven brain damage. In this way, we build on previous findings from our cohort in supporting the timepoint-based concept that burden of SE is detrimentally modulated by a longer SE duration and, to a lesser degree, by bilateral tonic-clonic semiology and reduced level of consciousness at initial clinical evaluation. In addition, location of PMA on baseline DWI/FLAIR was associated with distinct trajectories of deep grey matter volumes, with hippocampal PMA predicting hippocampal-thalamic atrophy and pulvinar PMA indicating a broader limbic-network pattern of damage.

MRI-based measures of grey matter volume and cortical thickness provide *in vivo* macrostructural proxies of grey matter integrity.^37^ These metrics are sensitive to underlying cytoarchitectural change and reflect neurodegenerative processes.^38^ The impact of burden model comprises the cumulative damage driven by prolonged seizure activity in the brain and the rest of the body.^14^ Current understanding of SE-induced brain damage is derived primarily from animal models, where prolonged seizures led to neuronal cell death, glial reactivity and tau accumulation.^39–42^ Evidence of epilepsy-driven brain injury in humans is scarce. A serial imaging study in 19 individuals with refractory SE demonstrated a reduction of the ventricular brain ratio as a marker for global brain shrinkage.^43^ In a paediatric population with febrile SE, the Consequences of Prolonged Febrile Seizures in Childhood (FEBSTAT) trial linked hippocampal T2 hyperintensity shortly after SE with development of hippocampal sclerosis and epilepsy over a 10-year follow-up period.^9^ Blood biomarker studies in humans further support this concept, with elevations in circulating markers of neuronal and glial damage such as neuron-specific enolase and neurofilament light chain reported during or shortly after SE.^44–47^

In our study, we report a tendency for progressive cortical thinning involving medial frontal and parietal areas over time. This contrasts with unexpected volume gain of several subcortical structures, such as the accumbens, putamen and caudate nucleus. This finding was consistent across most SE participants and is yet of unclear cause, warranting long-term follow-up to evaluate for transient swelling as well as external validation. An effect of aetiology is unlikely, as it did not seem to independently affect the rate of subcortical volume change in our SE cohort. We hypothesize this relates to middle-term network-remodelling involving key grey matter nuclei after SE. Among these, the thalamus is a core structure of the epileptogenic network and has been implicated in seizure-driven damage.^48,49^ In our SE cohort, thalamic volume tended to increase over time, although this change did not differ significantly from normal aging or chronic drug-resistant epilepsy. Even so, its rate of volume gain was positively and independently influenced by a convulsive semiology, likely reflecting the toll of sustained bilateral tonic-clonic activity on large-scale network reorganisation.^50^ Occurrence of PMA in the pulvinar nucleus of the thalamus, however, seemed to counteract thalamic volume gain bilaterally.

The most consistent feature associated with SE was hippocampal atrophy. In our cohort, a single episode of SE was associated with a faster rate of hippocampal atrophy than the average rate observed in drug-resistant focal epilepsy. This finding suggests that the predominant structural impact in epilepsy may be driven by the early injury itself, rather than by subsequent ongoing (i.e. self-limited) seizures, a concept that aligns with previous neuropsychological evidence.^51^ In regard to the vulnerability of the hippocampus during the peri-ictal state, our findings are also supported by a substantial body of imaging and neuropathological evidence. Previous neuroimaging data have consistently indicated a particular vulnerability of the CA1 hippocampal subfield to peri-ictal states, where glutamate-mediated excitotoxicity promotes cytotoxic oedema and leads to irreversible tissue damage and tau phosphorylation.^41,52–55^ The observed trends of grey matter atrophy affecting other limbic areas, such as the parahippocampal gyrus and cingulum are possibly secondary to primary hippocampal atrophy. In addition, longer duration of SE and a convulsive semiology also contributed independently to bilateral hippocampal atrophy in our patient collective. In this context, our findings build on previous observations by delivering a more precise quantification and prediction of mid-to long-term hippocampus atrophy following SE.^8,9^

In our statistical model, SE emerged as the strongest predictor of brain atrophy. According to our findings, the longer the duration of SE, the steeper the cortical atrophy in widespread neocortical areas as well as hippocampal volume decline. Reduced level of consciousness on admission, on the other hand, predicted medial frontal and parietal cortical thinning, without implicating subcortical grey matter. Both these features are known markers of more extensive network engagement during SE, as observed in connectivity studies in temporal lobe epilepsy, where impaired consciousness in focal seizures was linked to long-range network effects via thalamo-cortical desynchronization.^50,56,57^ Clinical and epidemiological studies further strengthen the link between impaired consciousness on initial clinical evaluation and subsequent brain damage, as decreased consciousness was associated with higher mortality, particularly in NCSE with coma.^58,59^ After adjustment for SE duration and aetiology, CSE was associated with a markedly faster rate of bilateral hippocampal atrophy than both drug-resistant focal epilepsy and healthy controls, representing the steepest volume decline across the three semiologies. Together with the pronounced thinning observed in medial cortical areas, this indicates a greater structural burden than that of either NCSE other PM-SE. Nevertheless, we also reinforce growing view that NCSE leads to substantial brain injury, independently of its typically longer duration. ^15,60^ In our cohort, NCSE predicted a rate of hippocampal atrophy approximately four-fold greater than that observed in healthy controls or in drug-resistant focal epilepsy.

The presence of PMA in DWI/FLAIR imaging has been suggested as a surrogate marker of t2, that is, the time after SE onset beyond which brain damage may be established.^11^ Here, occurrence of PMA in the hippocampus at baseline imaging was associated with a subsequent atrophy pattern involving the ipsilateral hippocampus, thalamus and amygdala as compared to absence of hippocampal PMA, a finding that is plausible given the well described hippocampo-thalamic connectivity in epilepsy.^61,62^ This interpretation is further supported by recent evidence from our cohort that PMA on DWI/FLAIR imaging were independently linked to a cumulative seizure risk exceeding 60%, consistent with the idea that hippocampal injury may lead to post-SE epileptogenesis, possibly via hippocampus sclerosis.^63^ By contrast, pulvinar PMA signaled a more distributed pattern of medium-term damage, predicting bilateral thalamic and amygdalar atrophy and thus suggesting involvement of a broader limbic network rather than a predominantly mesiotemporal process. This notion is emphasized by the fact that all 5 cases of pulvinar PMA were unilateral and bilateral involvement was observed. Despite the close connectivity between the pulvinar and several cortical regions, we did not observe a significant cortex involvement.^64,65^ Cortical PMA, in turn, was associated with widespread longitudinal volume changes across deep grey matter nuclei, including ipsilateral hippocampal atrophy, once again arguing for network-level consequences of acute peri-ictal cortical injury rather than isolated cortical damage alone. The absence of significant longitudinal cortical thickness changes following cortical PMA owes, most probably, to the broad lesion distribution of cortical lesions, which likely limits vertex-wise statistical power.

This study has several limitations. First, the retrospective study design inherently limits causal inference. Second, long-term follow-up data were not available, as the last MRI in the SE group was performed on average 5 months after SE, precluding assessment of longer-term brain changes, in particular volume gain recovery of deep grey matter structures. Third, because 81% of our SE participants exhibited PMA, the analysis is therefore biased toward more severe cases. Fourth, we did not account for the effects of prolonged sedation and treatment with potentially neurotoxic medications on cortical morphology. Fifth, small sample size does not allow subcategorisation of acute symptomatic aetiologies.^66^ Finally, limited baseline comparability between groups, substantial variability in interscan intervals, and the use of different MRI scanners despite application of harmonisation tools potentially introduced unwanted variability.

## Conclusion

Our findings suggest that sustained ictal activity leaves a measurable structural imprint on the brain that continues to evolve over months, beyond what would be expected from the underlying aetiology alone. Within a relatively short follow-up period, SE was associated with a distinct pattern of brain change characterised by progressive hippocampal atrophy, subtle yet spatially coherent cortical thinning of limbic structures, and subcortical volume increases. Importantly, these changes appear to be shaped the strongest by a longer duration of SE, thereby corroborating the timepoint-based concept of SE.^4^ To a lesser degree, brain structure is modulated by bilateral tonic-clonic activity and impaired consciousness on admission. Furthermore, a non-convulsive semiology was independently associated with hippocampal atrophy, emphasizing once again its damaging potential. Our observations reinforce the vulnerability of the hippocampus and limbic structures to ictal-driven brain damage and support the view that timely seizure control is critical for preserving functional and structural brain integrity.

## Data availability

The data supporting the findings of this study are available from the corresponding author upon reasonable request. Study datasets are not made publicly available due to ethical and data protection restrictions. Data pertaining the healthy control group was obtained on 2025-06-04 from the Parkinson’s Progression Markers Initiative (PPMI) database (www.ppmi-info.org/access-data-specimens/download-data), RRID:SCR_006431. For up-to-date information on the study, visit www.ppmi-info.org.

## Data Availability

Data regarding the status epilepticus drug-resistant focal epilepsy may be conceded upon reasonable request. Data pertaining the healthy control group was obtained on 2025-06-04 from the Parkinsons Progression Markers Initiative (PPMI) database (www.ppmi-info.org/access-data-specimens/download-data) RRID SCR_006431.

