## Supplementary Material for "Mapping the Cerebral Burden of Status Epilepticus – Results from a Longitudinal MRI Study"

[4.3 Per group slopes for FWE/FDR<0.05 clusters	13](#_Toc226551385)

### Supplementary Information 1: MRI protocols in SE and focal epilepsy

Individuals with SE were all scanned with the same MRI scanner. These data were acquired between February 2019 and April 2024 on a 3T MRI Philipps scanner. 3-dimensional (3D) T1 was obtained in a sagittal alignment with echo time (TE) 4.04 ms, repetition time (TR) 8.66 ms, field of view (FOV) 320 mm x 320 mm, and voxel size of 1x1x1 mm^3^.

MRI data in individuals with drug-resistant focal epilepsy were acquired with two different MRI scanners. All longitudinal scans per participant were obtained on a single scanner. Here, 25 individuals underwent the same protocol as the SE cohort, and in 9 participants MRI acquisition was undertaken on a Magnetom Prisma-Fit 3T MRI scanner using a 3D multi-echo magnetization prepared rapid gradient-echo imaging sequence (TE 2.2, TR 2.2 milliseconds, FOV 320 x 300 mm, voxel size of 0.8 x 0.8 x0.8 mm^3^).

### Supplementary Information 2: Criteria to define PMA

We defined diffusion-restricted lesions, FLAIR hyperintensity and hyperperfusion in ASL as PMA according to the following criteria:

1. Location of lesions did not respect defined vascular territories.
2. Lesions were in anatomically typical regions for PMA (cortex, hippocampus, amygdala, nucleus pulvinar of thalamus).^1^
3. Diffusion-restricted lesions were classified as PMA if quantitative analysis revealed a signal intensity ratio <1.495 on DWI and >0.735 on apparent diffusion coefficient.^2^
4. Lesions were not classified as PMA if they could be attributed to primary lesions of the CNS (e.g., brain tumors, old or acute ischemic stroke, old or acute cerebral hemorrhage, brain abscesses, encephalitis, etc.).

### Supplementary Information 3: Description of healthy volunteers

We analysed a total of 63 healthy volunteers from the Parkinson Progression Marker Initiative (PPMI) dataset.^3^

Scans used for this longitudinal cohort were acquired on 3T Siemens MRI scanners (Siemens Medical, Erlangen, Germany). A magnetization-prepared rapid gradient echo (MPRAGE) sequence was used to acquire high-resolution T1-weighted anatomical images (repetition time=2300 ms, echo time=2.98 ms, flip angle=9°, 240 x 256 matrix, 160-192 slices, slice thickness=1.0 mm, voxel size=1x1x1mm^3^).

The T1 acquisition protocol followed ADNI-3 sequence parameter recommendations: http://adni.loni.usc.edu/wp-content/uploads/2017/07/ADNI3-MRI-protocols.pdf

Detailed description of MR-acquisition protocol can be found in the PPMI MRI Technical Operations Manual: http://www.ppmi-info.org/wp-content/uploads/2017/06/PPMI-MRI-Operations-Manual-V7.pdf

### Supplementary Table 1: Global measures of brain structure

|  | **Change in**  **ml/month** | **CI 95%** | **T value** | **P value** |
| --- | --- | --- | --- | --- |
| **Healthy controls** |  |  |  |  |
| GMV | -0.156 | -0.897 0.584 | -0.503 | 0.631 |
| WMV | 0.121 | -0.434 0.676 | 0.529 | 0.632 |
| **Focal epilepsy** |  |  |  |  |
| GMV | -0.214 | -0.922 0.494 | -0.598 | 0.551 |
| WMV | 0.083 | -0.445 0.612 | 0.312 | 0.755 |
| **SE** |  |  |  |  |
| GMV | -0.814 | -1.881 0.253 | -1.593 | 0.127 |
| WMV | -0.812 | -1.592 -0.033 | -2.165 | 0.042 |
|  | **Difference in**  **ml/month** | **CI 95%** | **T value** | **P value** |
| **SE vs. healthy controls** |  |  |  |  |
| GMV | -0.600 | -1.865 0.666 | -0.959 | 0.343 |
| WMV | -0.896 | -1.824 0.032 | -1.950 | 0.049 |
| **SE vs. focal epilepsy** |  |  |  |  |
| GMV | -0.658 | -1.938 0.623 | -1.098 | 0.290 |
| WMV | -0.933 | -1.869 0.003 | -2.127 | 0.050 |

Abbreviations: CI, confidence interval; GMV: grey matter volume; WMV: white matter volume

### Supplementary Table 2: Per-group rate of longitudinal change

| **Cortical thickness** | | |  |  |  |
| --- | --- | --- | --- | --- | --- |
|  | **Side** | **ROI** | **No. vertices** | **Mean T-value** | ***P*_uncorrected_** |
| **Healthy controls** | | |  |  |  |
|  | RH | fusiform | 11 | -2.03 | <0.01 |
|  | RH | parahippocampal | 30 | -1.97 | <0.05 |
| **Focal epilepsy** | |  |  |  |  |
|  | LH | lateraloccipital | 47 | -2.97 | <0.01 |
|  | LH | parahippocampal | 33 | -2.9 | <0.01 |
|  | RH | parahippocampal | 45 | -3.6 | <0.01 |
|  | RH | posteriorcingulate | 58 | 3.51 | <0.01 |
|  | RH | precuneus | 15 | -2.1 | <0.05 |
|  | RH | lateraloccipital | 13 | -2.11 | <0.05 |
| **SE** | |  |  |  |  |
|  | LH | superiortemporal | 65 | -3.4 | <0.01 |
|  | LH | isthmuscingulate | 20 | -2.33 | <0.05 |
|  | LH | fusiform | 13 | 2.2 | <0.05 |
|  | LH | superiorfrontal | 10 | -2.54 | <0.01 |
|  | LH | parahippocampal | 23 | -3.92 | <0.001 |
|  | RH | posteriorcingulate | 101 | -3.08 | <0.01 |
|  | RH | superiorparietal | 31 | 3.11 | <0.01 |
|  | RH | precuneus | 22 | 2.44 | <0.05 |
|  | RH | parahippocampal | 11 | -2.15 | <0.05 |
|  | RH | entorhinal | 10 | -2.39 | <0.05 |

| **Subcortical Volumes** | |  |  |  |  |
| --- | --- | --- | --- | --- | --- |
|  | **Side** | **ROI** | **Slope (mm^3^/month)** | **95% CI** | ***P*_FDR_** |
| **Healthy controls** |  |  |  |  |  |
|  | left | accumbens | 2.671 | [1.157, 4.185] | <0.05 |
|  | left | amygdala | -0.366 | [-5.716, 4.984] | .890 |
|  | left | caudate | 20.593 | [9.017, 32.168] | <0.05 |
|  | left | hippocampus | -12.911 | [-18.185, -7.638] | .001 |
|  | left | pallidum | 2.068 | [-4.261, 8.398] | .571 |
|  | left | putamen | 20.185 | [6.991, 33.378] | <0.01 |
|  | left | thalamus | 13.256 | [-4.843, 31.355] | .205 |
|  | right | accumbens | 2.827 | [1.334, 4.321] | <0.01 |
|  | right | amygdala | -5.299 | [-11.030, 0.432] | 0.103 |
|  | right | caudate | 18.585 | [7.287, 29.883] | <0.01 |
|  | right | hippocampus | -11.568 | [-15.925, -7.210] | <0.001 |
|  | right | pallidum | -2.428 | [-10.204, 5.349] | 0.571 |
|  | right | putamen | 18.706 | [4.734, 32.679] | <0.05 |
|  | right | thalamus | 11.467 | [-6.694, 29.628] | 0.261 |
| **Focal epilepsy** |  |  |  |  |  |
|  | left | accumbens | 0.612 | [-1.841, 3.066] | 0.759 |
|  | left | amygdala | 3.551 | [-2.345, 9.446] | 0.649 |
|  | left | caudate | 5.270 | [-8.021, 18.561] | 0.694 |
|  | left | hippocampus | -9.063 | [-26.776, 8.649] | 0.649 |
|  | left | pallidum | 10.755 | [-0.099, 21.609] | 0.505 |
|  | left | putamen | 10.114 | [-5.804, 26.032] | 0.649 |
|  | left | thalamus | 3.684 | [-20.577, 27.946] | 0.763 |
|  | right | accumbens | 0.491 | [-2.122, 3.104] | 0.759 |
|  | right | amygdala | 1.992 | [-3.224, 7.207] | 0.694 |
|  | right | caudate | 6.698 | [-6.384, 19.780] | 0.649 |
|  | right | hippocampus | -5.847 | [-24.359, 12.665] | 0.744 |
|  | right | pallidum | 9.997 | [-0.983, 20.977] | 0.505 |
|  | right | putamen | 7.992 | [-8.270, 24.254] | 0.649 |
|  | right | thalamus | 4.943 | [-19.196, 29.082] | 0.759 |

| **SE** |  |  |  |  |  |
| --- | --- | --- | --- | --- | --- |
|  | left | accumbens | 12.142 | [4.292, 19.991] | <0.05 |
|  | left | amygdala | 0.800 | [-6.498, 8.097] | 0.892 |
|  | left | caudate | 58.039 | [8.303, 107.774] | <0.05 |
|  | left | hippocampus | -120.244 | [-199.904, -40.585] | <0.05 |
|  | left | pallidum | 45.331 | [10.457, 80.206] | 0.435 |
|  | left | putamen | 95.833 | [33.627, 158.039] | <0.05 |
|  | left | thalamus | 100.161 | [24.168, 176.153] | <0.05 |
|  | right | accumbens | 12.141 | [2.539, 21.742] | <0.05 |
|  | right | amygdala | 0.526 | [-9.727, 10.779] | 0.919 |
|  | right | caudate | 72.813 | [13.943, 131.684] | <0.05 |
|  | right | hippocampus | -114.564 | [-185.011, -44.118] | <0.05 |
|  | right | pallidum | 40.858 | [2.735, 78.980] | <0.05 |
|  | right | putamen | 83.172 | [20.511, 145.833] | <0.05 |
|  | right | thalamus | 109.260 | [29.507, 189.012] | <0.05 |

### Supplementary Table 3: Group comparisons

#### 3.1 Left-right comparisons

| **Cortical thickness** | | |  |  |  |
| --- | --- | --- | --- | --- | --- |
|  | **Side** | **ROI** | **No. vertices** | **Mean T-value** | ***P*_uncorrected_** |
| **SE vs.**  **healthy controls** | | |  |  |  |
|  | RH | superiortemporal | 11 | -2.59 | <0.01 |
|  | RH | isthmuscingulate | 3 | -2.03 | <0.05 |
|  | RH | superiorfrontal | 1 | -2.05 | <0.05 |
|  | LH | parahippocampal | 23 | -2.50 | <0.01 |
|  | LH | superiorparietal | 18 | 2.57 | <0.001 |
| **SE vs.**  **Focal epilepsy** | | |  |  |  |
|  | LH | superiortemporal | 60 | -3.15 | <0.001 |
|  | LH | lateraloccipital | 44 | 2.57 | <0.001 |
|  | LH | fusiform | 33 | 2.52 | <0.01 |
|  | LH | supramarginal | 10 | 2.26 | <0.05 |
|  | LH | superiorfrontal | 9 | -2.5 | <0.01 |
|  | LH | isthmuscingulate | 7 | -2.15 | <0.05 |
|  | LH | rostralmiddlefrontal | 5 | -2.27 | <0.05 |
|  | RH | posteriorcingulate | 102 | -3.7 | <0.001 |
|  | RH | precuneus | 37 | 2.5 | <0.01 |
|  | RH | superiorparietal | 23 | 2.56 | <0.05 |
|  | RH | inferiorparietal | 17 | -2.35 | <0.05 |
|  | RH | caudalanteriorcingulate | 14 | -2.14 | <0.05 |
|  | RH | pericalcarine | 14 | 2.17 | <0.05 |
|  | RH | precentral | 13 | 2.21 | <0.05 |
|  | RH | lingual | 12 | 2.19 | <0.05 |
|  | RH | inferiortemporal | 10 | 2.17 | <0.05 |
|  | RH | entorhinal | 3 | -2.05 | <0.05 |

| **Subcortical Volumes** | |  |  |  |  |
| --- | --- | --- | --- | --- | --- |
|  | **Side** | **ROI** | **Slope difference (mm^3^/month)** | **T-value** | ***P*_FDR_** |
| **SE vs. healthy controls** | |  |  |  |  |
|  | left | accumbens | 5.05 | 2.4 | <0.05 |
|  | left | amygdala | 3.98 | 0.81 | 0.42 |
|  | left | caudate | 23.01 | 1.45 | 0.21 |
|  | left | hippocampus | -73.69 | -3.5 | <0.001 |
|  | left | pallidum | 36.14 | 3.08 | <0.05 |
|  | left | putamen | 34.37 | 2.39 | <0.05 |
|  | left | thalamus | 27.72 | 1.32 | .22 |
|  | right | accumbens | 5.27 | 2.2 | 0.07 |
|  | right | amygdala | 5.86 | 1.03 | 0.31 |
|  | right | caudate | 29.71 | 1.96 | 0.09 |
|  | right | hippocampus | -76.1 | -3.84 | <0.001 |
|  | right | pallidum | 45.81 | 1.61 | 0.15 |
|  | right | putamen | 41.59 | 2.59 | 0.04 |
|  | right | thalamus | 30.49 | 1.45 | 0.17 |
|  | right | accumbens | 5.27 | 2.2 | 0.07 |
|  | right | amygdala | 5.86 | 1.03 | 0.31 |
| **SE vs. focal epilepsy** | |  |  |  |  |
|  | left | accumbens | 7.22 | 3.55 | <0.001 |
|  | left | amygdala | 1.69 | 0.38 | 0.703 |
|  | left | caudate | 42.9 | 2.6 | <0.05 |
|  | left | hippocampus | -83.17 | -3.76 | <0.001 |
|  | left | pallidum | 34.11 | 2.87 | <0.01 |
|  | left | putamen | 46.43 | 3.46 | 0.001 |
|  | left | thalamus | 36.64 | 1.93 | <0.05 |
|  | right | accumbens | 7.67 | 3.19 | <0.01 |
|  | right | amygdala | -0.48 | -0.1 | 0.923 |
|  | right | caudate | 44.5 | 2.89 | <0.01 |
|  | right | hippocampus | -87.37 | -4.28 | <0.001 |
|  | right | pallidum | 77.26 | 2.59 | <0.05 |
|  | right | putamen | 56.45 | 3.62 | 0.001 |
|  | right | thalamus | 36.65 | 1.92 | 0.065 |

#### 3.2 Ipsi-contralateral comparisons

| **Cortical thickness** | | |  |  |  |
| --- | --- | --- | --- | --- | --- |
|  | **Side** | **ROI** | **No. vertices** | **Mean T-value** | ***P*_uncorrected_** |
| **SE vs. Focal epilepsy** | | |  |  |  |
|  | ipsi | posteriorcingulate | 56 | -3.72 | <0.001 |
|  | ipsi | precuneus | 54 | -2.58 | <0.01 |
|  | ipsi | superiortemporal | 46 | -2.76 | <0.01 |
|  | ipsi | entorhinal | 17 | -2.64 | <0.01 |
|  | ipsi | middletemporal | 8 | -2.29 | <0.05 |
|  | ipsi | fusiform | 4 | 2.19 | <0.05 |
|  | ipsi | lingual | 3 | -2.09 | <0.05 |
|  | ipsi | superiorfrontal | 2 | -2.2 | <0.05 |
|  | contra | inferiortemporal | 9 | 2.08 | <0.05 |
|  | contra | postcentral | 3 | 2.01 | <0.05 |

| **Subcortical Volumes** | |  |  |  |  |
| --- | --- | --- | --- | --- | --- |
|  | **Side** | **ROI** | **Slope difference (mm^3^/month)** | **T-value** | ***P*_FDR_** |
| **SE vs. focal epilepsy** | |  |  |  |  |
|  | ipsilateral | accumbens | 7.91 | 2.96 | <0.01 |
|  | ipsilateral | amygdala | -3.07 | -0.51 | 0.61 |
|  | ipsilateral | caudate | 71.94 | 2.6 | <0.05 |
|  | ipsilateral | hippocampus | -126.07 | -3.65 | <0.001 |
|  | ipsilateral | pallidum | 60.13 | 1.58 | 0.14 |
|  | ipsilateral | putamen | 48.51 | 3.03 | <0.01 |
|  | ipsilateral | thalamus | 35.45 | 1.67 | 0.14 |
|  | contralateral | accumbens | 18.75 | 2.6 | <0.05 |
|  | contralateral | amygdala | 4.21 | 1.17 | 0.28 |
|  | contralateral | caudate | 94.94 | 2.67 | <0.05 |
|  | contralateral | hippocampus | -34.66 | -0.65 | 0.52 |
|  | contralateral | pallidum | 81.9 | 2.16 | 0.06 |
|  | contralateral | putamen | 127.53 | 2.3 | <0.05 |
|  | contralateral | thalamus | 41.02 | 1.99 | 0.07 |

### Supplementary Figure 1

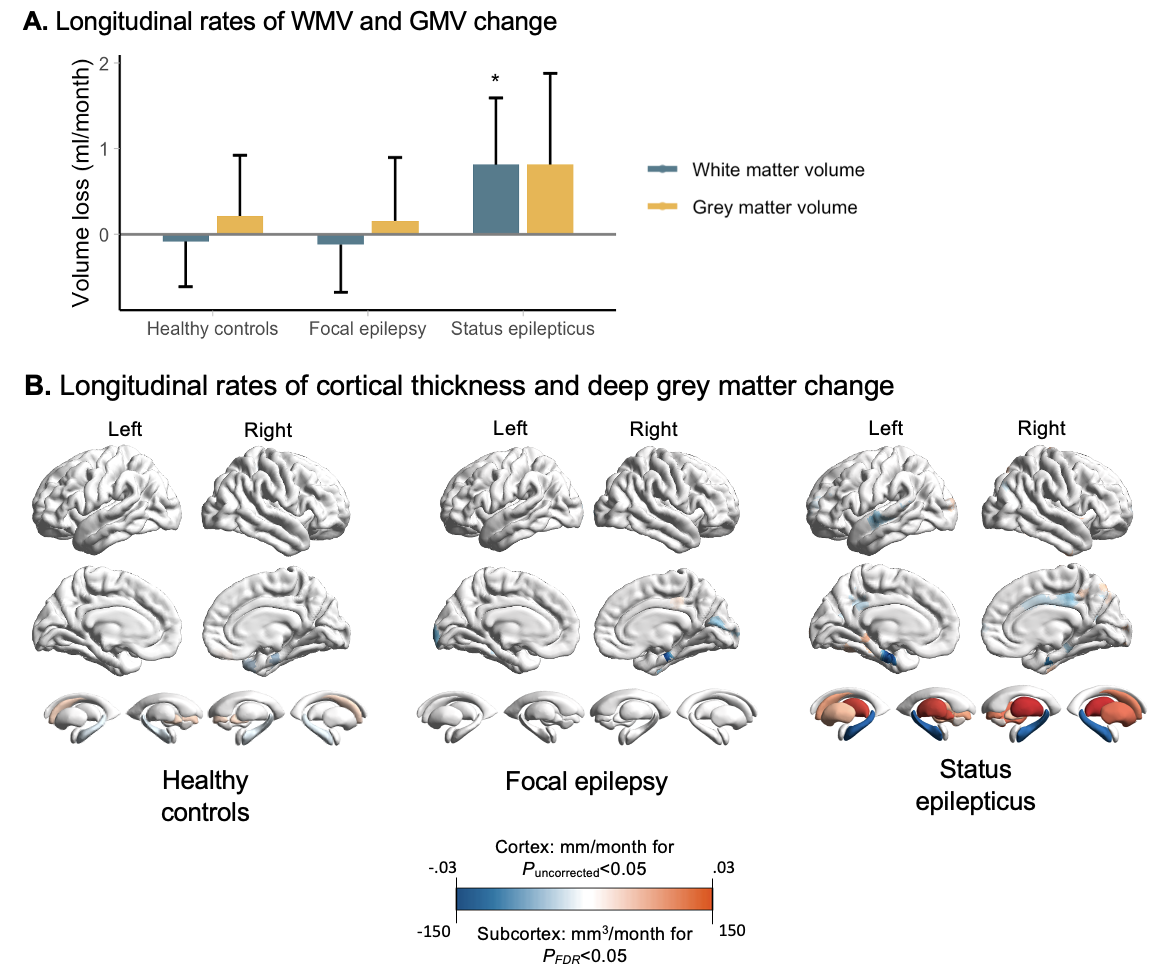

**Figure 1 Per-group progressive brain changes** (A) Global brain volume loss, including WMV and GMV. Volume loss was calculated via subtraction of parameter estimates at baseline imaging from those at the last MRI follow-up and averaged across each group. (B) Whole-brain plots display per-group longitudinal rates of change of cortical thickness (mm/month) and subcortical structure volume (mm^3^/month). No cortical clusters survived RFT correction at family-wise error (FWE) <0.05, therefore clusters with uncorrected *P* values <0.05 are displayed. For subcortical volumes, results are restricted to structures with false discovery rate correction (FDR) at *P*<0.05.

Abbreviations: FWE, family-wise error; RFT, random field theory; WMV, white matter volume; GMV, grey matter volume.

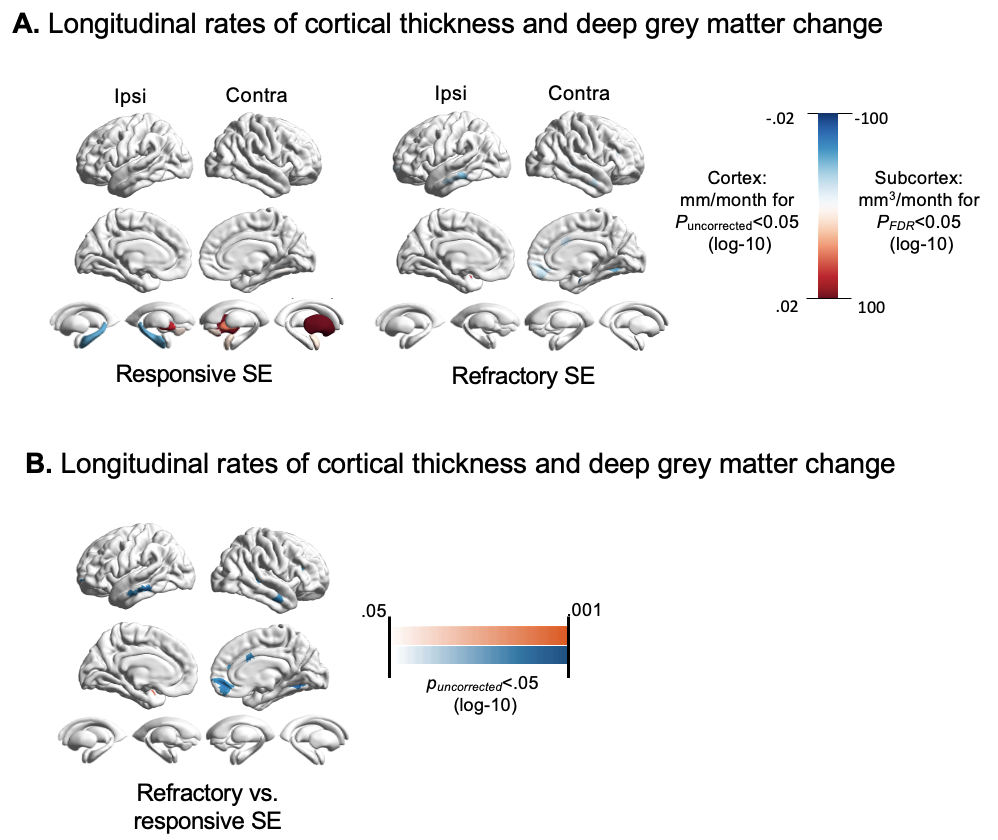

### Supplementary Figure 2

**Supplementary** **figure 2 Longitudinal brain changes according to drug responsiveness of SE** (A) Whole-brain plots display per-group longitudinal rate of change of cortical thickness (mm/month) and subcortical structure volume (mm^3^/month). No cortical clusters survived RFT correction at family-wise error (FWE) <0.05, therefore clusters with uncorrected *P* values <0.05 are displayed. For subcortical volumes, results are restricted to structures with false discovery rate correction (FDR) at *P*<0.05. In (B), brain-wide maps show the longitudinal slope differences between drug-refractory and drug-responsive SE.

### Supplementary Figure 3

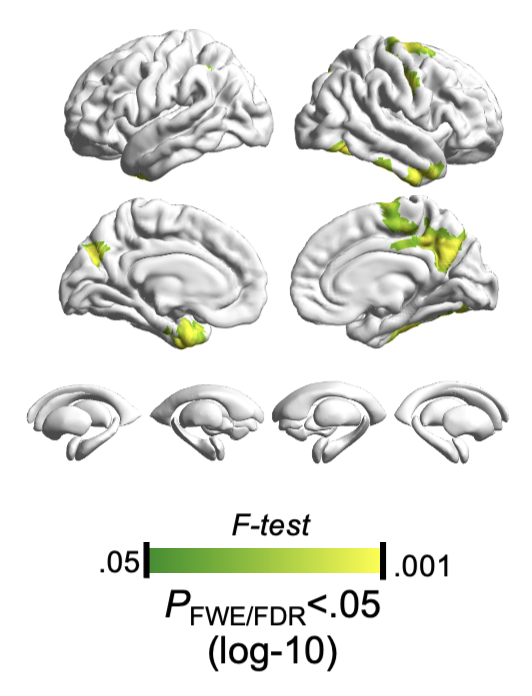

**Supplementary figure 3 Longitudinal variance of cortical thickness explained by aetiology**

Cortical effects were identified using omnibus tests of the aetiology by time in models and are shown at RFT-corrected *P_FWE_*<0.05. Subcortical effects were tested in analogous mutually adjusted mixed-effects models and are shown at FDR-corrected *P*<0.05.

### Supplementary Table 4: Modulators of brain atrophy

#### 4.1 Independent effects on cortical thickness

| ***Level of consciousness*** | |  |  |  |
| --- | --- | --- | --- | --- |
| **Side** | **No. Vertices** | **Cluster *P*_FWE_** | **Peak F** | **ROIs** |
| ipsi | 174 | <0.05 | 21.81 | paracentral (62.6%, n=109); posteriorcingulate (23.6%, n=41); superiorfrontal (13.8%, n=24) |
| contra | 414 | <0.01 | 23.47 | precuneus (43.0%, n=178);  paracentral (31.6%, n=131); posteriorcingulate (15.0%, n=62); superiorfrontal (9.9%, n=41); isthmuscingulate (0.5%, n=2) |
| contra | 179 | <0.01 | 36.33 | fusiform (70.9%, n=127);  lingual (16.2%, n=29);  lateraloccipital (8.4%, n=15); inferiortemporal (4.5%, n=8) |

| ***Semiology*** | |  |  |  |
| --- | --- | --- | --- | --- |
| **Side** | **No. Vertices** | **Cluster *P*_FWE_** | **Peak F** | **ROIs** |
| contra | 354 | <0.0001 | 20.58 | fusiform (56.9%, n=193); parahippocampal (26.0%, n=88); lingual (12.7%, n=43);  lateraloccipital (1.8%, n=6);  entorhinal (1.5%, n=5);  inferiortemporal (1.2%, n=4) |
| contra | 338 | <0.01 | 9.86 | precuneus (64.1%, n=211); isthmuscingulate (22.8%, n=75); posteriorcingulate (13.1%, n=43) |

| ***Duration of SE*** | |  |  |  |
| --- | --- | --- | --- | --- |
| **Side** | **No. Vertices** | **Cluster pFWE** | **Peak T** | **ROIs** |
|  | 246 | <0.0001 | -33.37 | precuneus (74.4%, n=183);  cuneus (14.2%, n=35);  superiorparietal (11.4%, n=28) |
| ipsi | 228 | <0.0001 | -24.31 | supramarginal (86.0%, n=196); inferiorparietal (14.0%, n=32) |
| ipsi | 138 | <0.001 | -20.28 | fusiform (30.2%, n=35); temporalpole (27.6%, n=32); entorhinal (23.3%, n=27);  superiortemporal (10.3%, n=12); inferiortemporal (8.6%, n=10) |
| ipsi | 263 | <0.001 | -15.73 | paracentral (40.7%, n=107);  superiorfrontal (37.6%, n=99);  precentral (21.7%, n=57) |
| contra | 1773 | <0.001 | -21.75 | superiorfrontal (27.9%, n=494);  precentral (20.2%, n=359);  precuneus (20.2%, n=358);  paracentral (11.3%, n=200); rostralmiddlefrontal (8.8%, n=156); postcentral (3.4%, n=61);  posteriorcingulate (2.5%, n=45); caudalmiddlefrontal (1.9%, n=33);  cuneus (1.8%, n=32);  superiorparietal (1.1%, n=19); isthmuscingulate (0.9%, n=16) |
| contra | 544 | <0.0001 | -42.72 | fusiform (41.4%, n=225);  inferiortemporal (21.3%, n=116); middletemporal (16.4%, n=89); parahippocampal (7.2%, n=39);  lingual (7.2%, n=39);  lateraloccipital (3.5%, n=19); superiortemporal (3.1%, n=17) |

#### 4.2 Independent effects on subcortical volumes

| ***Semiology*** |  |  |  |
| --- | --- | --- | --- |
| ***Side*** | **ROI** | **F-value** | ***P*_FDR_** |
| Ipsi | Hippocampus | 9.87 | <0.05 |
| Ipsi | Thalamus | 10.42 | <0.05 |
| Contra | Hippocampus | 16.02 | <0.05 |
| Contra | Thalamus | 9.14 | <0.05 |

| ***Duration of SE*** |  |  |  |
| --- | --- | --- | --- |
| ***Side*** | **ROI** | **T-value** | ***P*_FDR_** |
| Ipsi | Hippocampus | -9.10 | <0.01 |
| Contra | Hippocampus | -10.10 | <0.05 |

#### 4.3 Per group slopes for *P*_FWE/FDR_<0.05 clusters

| ***Semiology*** | **Difference in mm/mm^3^ per month** | | **95% CI** | **T value** | ***P* value** |
| --- | --- | --- | --- | --- | --- |
| **Cortical thickness** |  | |  |  |  |
| CSE | -0.19 | | [-0.21, -0.17] | -17.30 | <0.01 |
| other PM-SE | 0.08 | | [0.01, 0.02] | 3.79 | <0.01 |
| NCSE | 0.00 | | [-0.01, 0.02] | 0.11 | .91 |
| **Ipsilateral hippocampus** | | |  |  |  |
| CSE | -370.56 | | [-457.30, -283.82] | -8.51 | <0.01 |
| other PM-SE | -136.38 | | [-169.33, -103.44] | -8.25 | <0.01 |
| NCSE | -55.31 | | [-73.97, -36.64] | -5.9 | <0.01 |
| **Contralateral hippocampus** | | |  |  |  |
| CSE | | -480.03 | [-526.00, -434.05] | -20.82 | <0.01 |
| other PM-SE | | -102.16 | [-152.26, -117.19] | -15.32 | <0.01 |
| NCSE | | -343.48 | [-55.66, -35.69] | -9.12 | <0.01 |
| ***Consciousness level*** | **Difference in mm/month** | |  | **T value** | ***P* value** |
| **Cortical thickness** | |  |  |  |  |
| Alert/somnolent | | 0 | [-0.14, 0.13] | -0.02 | 0.99 |
| Stupor/comatose | | -0.20 | [-0.28, 0.12] | -5.07 | <0.01 |

#### 4.4 Group comparisons for semiology and consciousness level

| ***Semiology*** | **Difference in mm or mm^3^ per month** | **95% CI** | **T value** | **P value** |
| --- | --- | --- | --- | --- |
| **Cortical thickness** |  |  |  |  |
| CSE vs. other PM-SE | -0.20 | [-0.23, -0.18] | -17.51 | <0.01 |
| CSE vs. NCSE | -0.19 | [-0.21, -0.17] | -16.93 | <0.01 |
| **Ipsilateral hippocampus** | |  |  |  |
| CSE vs other PM-SE | -234.18 | [-326.96, -141.39] | -5.93 | <0.01 |
| CSE vs NCSE | -315.25 | [-403.98, -226.53] | -7.08 | <0.01 |
| **Contralateral hippocampus** | |  |  |  |
| CSE vs other PM-SE | -345.30 | [-394.50, -296.10] | -13.99 | <0.01 |
| CSE vs NCSE | -434.35 | [-481.39, -387.30] | -18.41 | <0.01 |

|  |  |  |  |  |
| --- | --- | --- | --- | --- |
| ***Consciousness level*** | **Difference in mm/month** | **95% CI** | **T value** | **P value** |
| **Cortical thickness** |  |  |  |  |
| Coma vs Alert | -0.20 | [-0.04, -0.36] | 2.55 | 0.01 |

#### 4.5 Slopes by different SE durations

| ***SE* *Duration*** | **Difference in mm or mm^3^ per month** | **CI 95%** | **T-value** | **P-value** |
| --- | --- | --- | --- | --- |
| **Cortical thickness** |  |  |  |  |
| 1 h | -0.1237 | [-0.2173, -0.03] | -2.631 | 0.010 |
| 5 h | -0.1863 | [-0.2591, -0.1136] | -5.102 | <0.001 |
| 10 h | -0.2209 | [-0.3056, -0.1363] | -5.201 | <0.001 |
| 30 h | -0.2800 | [-0.409, -0.1511] | -4.331 | <0.001 |
| **Ipsilateral hippocampus** | |  |  |  |
| 1 h | -411.17 | [-722.64, -99.71] | -2.64 | 0.010 |
| 5 h | -694.49 | [-941.31, -447.67] | -5.62 | <0.001 |
| 10 h | -850.80 | [-1126.78, -574.83] | -6.15 | <0.001 |
| 30 h | -1.118.00 | [-1518.16, -717.84] | -5.57 | <0.001 |
| **Contralateral hippocampus** | |  |  |  |
| 1 h | -91.44 | [-294.12, 111.25] | -0.90 | 0.371 |
| 5 h | -328.63 | [-495.89, -161.37] | -3.92 | <0.001 |
| 10 h | -459.50 | [-653.40, -265.60] | -4.72 | <0.001 |
| 30 h | -683.19 | [-969.12, -397.27] | -4.76 | <0.001 |

### Supplementary Table 5: Group comparisons between PMA status

|  | **Side** | **ROI** | **Slope difference (mm3/month)** | **T-value** | ***P*_FDR_** |
| --- | --- | --- | --- | --- | --- |
| **PMA hippocampus** | |  |  |  |  |
|  | ipsilateral | accumbens | -7.46 | -1.03 | 0.43 |
|  | ipsilateral | amygdala | -107.83 | -2.10 | <0.05 |
|  | ipsilateral | caudate | -41.55 | -1.05 | 0.43 |
|  | ipsilateral | hippocampus | -145.66 | -1.93 | <0.01 |
|  | ipsilateral | pallidum | -38.19 | -1.21 | 0.43 |
|  | ipsilateral | putamen | -31.44 | -0.69 | 0.57 |
|  | ipsilateral | thalamus | -160.36 | -2.21 | <0.01 |
|  | contralateral | accumbens | -1.94 | -0.25 | 0.86 |
|  | contralateral | amygdala | -19.27 | -1.42 | 0.42 |
|  | contralateral | caudate | -20.73 | -0.44 | 0.86 |
|  | contralateral | hippocampus | -71.91 | -1.35 | 0.42 |
|  | contralateral | pallidum | -56.98 | -1.59 | 0.42 |
|  | contralateral | putamen | -15.32 | -0.28 | 0.86 |
|  | contralateral | thalamus | 7.71 | 0.18 | 0.86 |
| **PMA Pulvinar** | |  |  |  |  |
|  | ipsilateral | accumbens | -14.21 | -3.89 | <0.001 |
|  | ipsilateral | amygdala | -50.09 | -2.55 | <0.05 |
|  | ipsilateral | caudate | -40.18 | -0.88 | 0.53 |
|  | ipsilateral | hippocampus | 3.36 | 0.11 | 0.92 |
|  | ipsilateral | pallidum | -23.41 | -0.52 | 0.70 |
|  | ipsilateral | putamen | -83.45 | -1.48 | 0.25 |
|  | ipsilateral | thalamus | -82.65 | -2.73 | <0.05 |
|  | contralateral | accumbens | -0.95 | -0.09 | 0.97 |
|  | contralateral | amygdala | -37.08 | -3.51 | <0.01 |
|  | contralateral | caudate | -27.17 | -0.44 | 0.93 |
|  | contralateral | hippocampus | 3.01 | 0.04 | 0.97 |
|  | contralateral | pallidum | -27.51 | -0.53 | 0.93 |
|  | contralateral | putamen | -38.36 | -0.51 | 0.93 |
|  | contralateral | thalamus | -110.48 | -3.40 | <0.001 |
| **PMA Cortex** | |  |  |  |  |
|  | ipsilateral | accumbens | 18.34 | 3.70 | 0.00 |
|  | ipsilateral | amygdala | -22.02 | -1.11 | 0.27 |
|  | ipsilateral | caudate | 67.56 | 2.55 | <0.05 |
|  | ipsilateral | hippocampus | -97.62 | -3.21 | 0.01 |
|  | ipsilateral | pallidum | 48.56 | 1.54 | 0.18 |
|  | ipsilateral | putamen | 78.97 | 1.96 | 0.09 |
|  | ipsilateral | thalamus | 48.15 | 1.39 | 0.20 |
|  | contralateral | accumbens | 20.51 | 3.16 | <0.05 |
|  | contralateral | amygdala | 21.12 | 2.42 | <0.05 |
|  | contralateral | caudate | 109.19 | 4.36 | <0.001 |
|  | contralateral | hippocampus | -31.64 | -0.56 | 0.58 |
|  | contralateral | pallidum | 53.82 | 1.46 | 0.17 |
|  | contralateral | putamen | 120.62 | 2.66 | <0.05 |
|  | contralateral | thalamus | 86.77 | 2.33 | <0.05 |
